# Association of childhood adversity with frailty and the mediating role of unhealthy lifestyle: Findings from the UK biobank

**DOI:** 10.1101/2023.02.08.23285634

**Authors:** Gan Yang, Xingqi Cao, Jie Yu, Xueqin Li, Liming Zhang, Jingyun Zhang, Chao Ma, Ning Zhang, Qingyun Lu, Chenkai Wu, Xi Chen, Emiel O. Hoogendijk, Thomas M. Gill, Zuyun Liu

**Affiliations:** School of Public Health and Second Affiliated Hospital, The Key Laboratory of Intelligent Preventive Medicine of Zhejiang Province, Zhejiang University School of Medicine, Hangzhou 310058, Zhejiang, China; School of Economics and Management, Southeast University, Nanjing 211189, Jiangsu, China; Department of Social Medicine School of Public Health and Center for Clinical Big Data and Analytics Second Affiliated Hospital, Zhejiang University School of Medicine, Hangzhou 310058, Zhejiang, China; School of Public Health, Nantong University, Nantong 226007, JiangSu, China; Duke Global Health Institute, Duke University, Durham, NC, USA; Department of Health Policy and Management, Yale School of Public Health, New Haven, CT 06520, United States of America; Department of Economics, Yale University, New Haven, CT 06520, United States of America; Department of Epidemiology & Data Science, Amsterdam Public Health research institute, Amsterdam UMC – location VU University medical center, Amsterdam, the Netherlands; Department of Internal Medicine, Yale School of Medicine, New Haven, CT 06520, United States of America

**Keywords:** Aging, Childhood adversity, Lifestyle, Frailty, Mediation analysis

## Abstract

**Background:** Childhood adversity and lifestyle have been associated with frailty in later life, but not much is known about factors that may explain these associations. An unhealthy lifestyle may play an important role in the pathway from childhood adversity to frailty. Therefore, this study aims to investigate the association of childhood adversity with frailty, and the mediating role of unhealthy lifestyle in the association.

**Methods:** This lifespan analysis included 152914 adults aged 40-69 years old from the UK Biobank. We measured childhood adversity with five items: physical neglect, emotional neglect, sexual abuse, physical abuse, and emotional abuse through online mental health survey. Frailty was measured by the frailty index; an unhealthy lifestyle score (range: 0-5) was calculated based on unhealthy body mass index, smoking, drinking, physical inactivity, and unhealthy diet at the baseline survey. Multiple logistic regression and mediation analysis were performed.

**Results:** A total of 10078 participants (6.6%) were defined as having frailty. Participants with any childhood adversity had higher odds of frailty. For example, in the fully adjusted model, with a one-point increase in cumulative score of childhood adversity, the odds of frailty increased by 41% (Odds Ratio: 1.41; 95% Confidence Interval: 1.39, 1.44). Unhealthy lifestyle partially mediated the associations of childhood adversity with frailty (mediation proportion: 4.4%-7.0%). The mediation proportions were largest for physical (8.2%) and sexual (8.1%) abuse.

**Conclusions:** Among this large sample, childhood adversity was positively associated with frailty, and unhealthy lifestyle partially mediated the association. This newly identified pathway highlights the potential of lifestyle intervention strategies among those who experienced childhood adversity (in particular, physical and sexual abuse) to promote healthy aging.

## Introduction

With population aging increasing worldwide, the promotion of healthy aging has gained more attention in recent years. A crucial component of healthy aging is the prevention of geriatric syndromes, such as frailty. The importance of the frailty concept, characterized by the decline of physiological systems and the ability to regulate stressors, is increasingly being acknowledged [1]. Additionally, frailty accounts for many adverse health outcomes such as disability, falls, hospitalization, and mortality, which brings a huge burden to society, caregivers, and families [2, 3].

Life course theory indicates that early exposure to environmental, psychosocial, and physical factors can influence the aging process [4, 5], frailty [6-14], and late-life health [15] (e.g. cardiovascular disease [16], mortality [17]). To date, five epidemiological studies have found a positive association between childhood adversity and frailty [18-22] while one study indicated a null association [23]. The conflicting findings might be partially due to the limited sample size. More importantly, not much is known about factors that may explain the childhood adversity-frailty association (if it was established) [22]. Campbell et al. [24] found that people who had adverse childhood experiences were more likely to be binge drinkers, heavy drinkers, smokers, or obese. Recent evidence also highlights that unfavorable lifestyle including lack of physical activity, poor nutrition, and sleep deprivation may synergistically influence health in later life [25]. Thus, we hypothesize that childhood adversity may influence frailty through unhealthy lifestyle, which, however, previous studies have not explored.

Therefore, this study aimed to examine the relationship between childhood adversity and frailty as well as the mediating role of unhealthy lifestyle in this association using data from approximately 150000 UK adults.

## Methods

### Study participants

This lifespan analysis used data from the UK Biobank (UKB), a large population-based cohort study with approximately half a million participants aged 40-69 years at recruitment in 2006-2010 [26]. UKB was approved by the North West-Haydock Research Ethics Committee and all participants electronically wrote informed consent at recruitment. A total of 156749 adults who were healthier, better educated, and had higher socioeconomic status than the overall biobank sample participated in both the baseline survey and the subsequent 2016 online mental health survey [27]. We excluded participants with missing data on childhood adversity (N=3728) and frailty (N=107), leaving 152914 participants in the analytic sample (**Figure S1**).

### Childhood adversity

Data on childhood adversity was sourced from the 2016 online mental health survey. Childhood adversity was characterized as physical neglect, emotional neglect, sexual abuse, physical abuse, and emotional abuse using the Childhood Trauma Screener [28] across five questions. Items were divided based on the options (“very often true”, “often true”, “sometimes true”, “rarely true”, “never true”), which were scored 0 or 1 according to the cut-off points shown in **Table S1** after inversely scoring two positive items (i.e., emotional neglect, physical neglect) [29]. The cumulative score of all five items ranged from 0 to 5, with a higher score indicating more childhood adversity. The items included are:

(1) Someone to take to doctor when needed as a child (physical neglect);

(2) Felt loved as a child (emotional neglect);

(3) Sexually molested as a child (sexual abuse);

(4) Physically abused by family as a child (physical abuse);

(5) Felt hated by family member as a child (emotional abuse).

### Frailty measures

Frailty was assessed at the baseline survey, using frailty index following the methods of Rockwood et al. [30] based on a series of indicators of ill-health in physiological and mental domains including diagnosed diseases, symptoms, and disabilities. The frailty index is well-validated for predicting mortality [2] and calculated through 49 self-reported items covering sensory, cranial, mental well-being, infirmity, cardiometabolic, musculoskeletal, immunological, cancer, pain, and gastrointestinal [31]. A frailty score, a continuous value ranging from 0 to 1, was obtained by dividing the number of deficits by the total number of deficits. For instance, a participant who has 10 items that scored 1 as opposed to 0 would have frailty scored 10/49 (i.e., 0.204). According to the literature [32], participants with a score over 0.21 were categorized as frailty, and those with a score less than 0.21 were categorized as non-frailty.

### Assessment of unhealthy lifestyle

An unhealthy lifestyle score was constructed according to five lifestyle factors (i.e., smoking status, alcohol consumption, diet, body mass index (BMI), and physical activity) sourced from structured questionnaires and 24 hours dietary recalls at the baseline survey.

Based on the recommendations of the World Health Organization, smoking more than 100 cigarettes in life was considered as unhealthy [33]. The daily consumption of one alcoholic drink or more for females and two drinks or more for males was considered as unhealthy [33]. For diet, having not achieved the intake goals for more than half of the following components: vegetables, fruits, whole grains, (shell)fish, vegetable oils, dairy products, unprocessed meats, sugar-sweetened beverages, and refined grains was considered as unhealthy [34]. A BMI out of the range of 18.5-24.9 kg/m^2^ was considered as unhealthy [35]. For physical activity, a) less than 150 minutes or 5 days per week of moderate physical activity; or b) less than 75 minutes or once per week of vigorous activity were considered as unhealthy levels of physical activity [36]. For each lifestyle factor, the unhealthy level was scored 1 and otherwise, was scored 0. The unhealthy lifestyle score ranged from 0 to 5 and a higher score indicated a higher level of unhealthy lifestyle.

### Covariates

The following covariates collected at baseline survey were included: chronological age, sex, ethnicity, educational level, occupation, Townsend deprivation index (TDI), and maternal smoking around birth. Ethnicity was classified as white, mixed, south Asian, black, Chinese, and others [37]. Educational level was defined as high (college or university degree), intermediate (A/AS levels or O levels/GCSEs or equivalent), and low (none of the aforementioned) [33]. The occupation was classified as working, retired, and others (unemployed, unpaid/voluntary work, full/part-time student, looking after home and/or family, unable to work because of sickness or disability, or did not answer) [38]. TDI consisted of data on housing, social class based on the postal code, and employment, with a higher TDI indicating a lower level of socioeconomic status [38].

### Statistical Analyses

The basic characteristics of the study participants were classed by frailty with mean±standard deviation (SD) for continuous variables and counts (percentage) for categorical variables. The basic characteristics across groups were compared by t-test for continuous variables and χ^2^ test for categorical variables, respectively. We imputed the missing values of smoking status (N=1632), alcohol consumption (N=16299), diet (N=1), physical activity (N=798), and covariates (N=20597) with the Multiple Imputation by Chained Equations [39], using five copies of imputation.

First, multiple logistic regression models were performed to estimate the associations between childhood adversity and frailty (**Figure 1**). We documented odds ratios (ORs) and corresponding 95% confidence intervals (CIs) from three models. Model 1 was adjusted for age and sex. Model 2 was additionally adjusted for ethnicity, occupation, educational level, TDI, and maternal smoking around birth. Model 3 was additionally adjusted for unhealthy lifestyle score.

**Figure 1.**
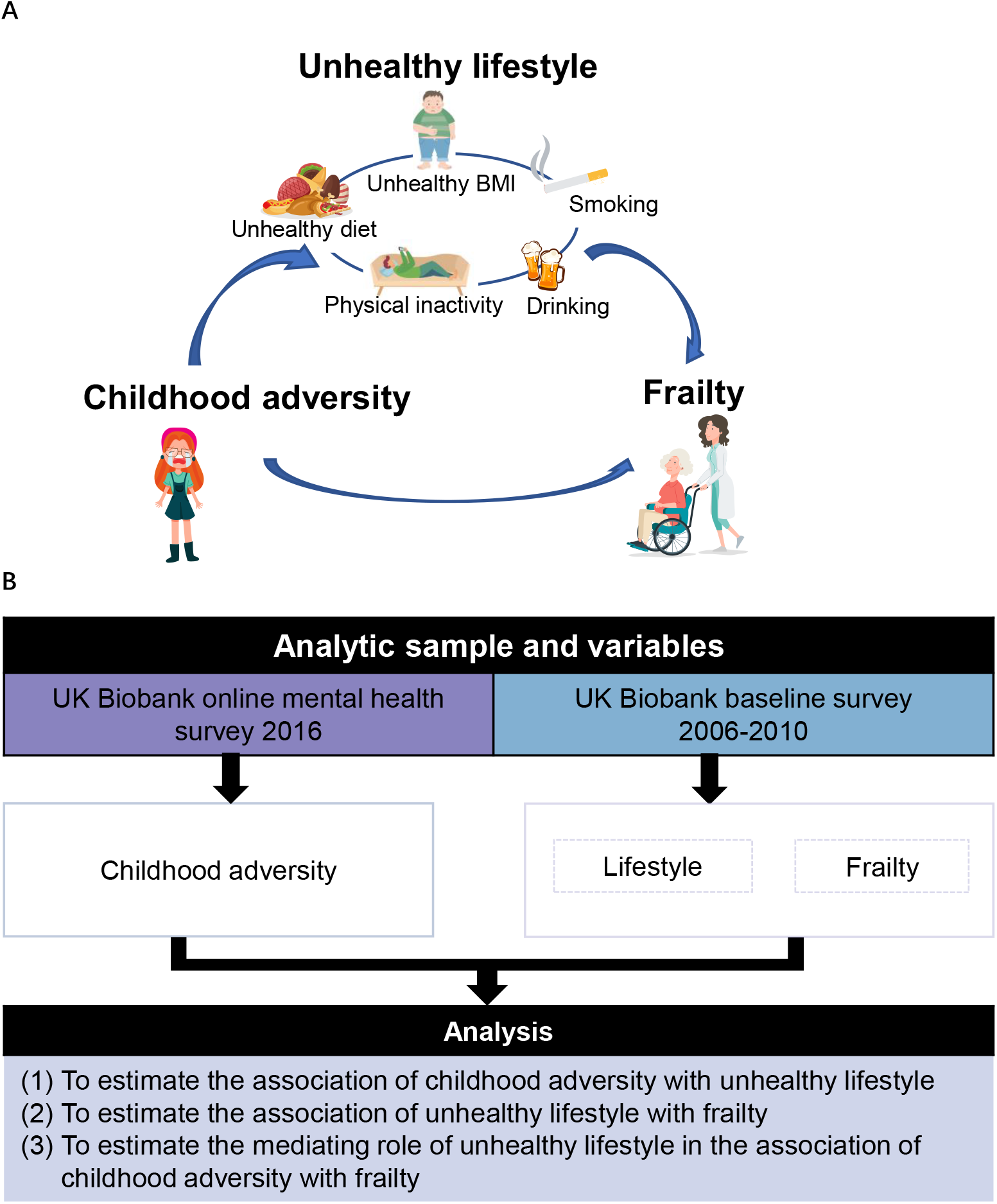
Roadmap for evaluating associations between childhood adversity, unhealthy lifestyle, and frailty.

Second, to examine whether unhealthy lifestyle mediates the associations of childhood adversity with frailty, we performed a mediation analysis. First, we used linear regression models to evaluate the associations of childhood adversity with unhealthy lifestyle score in two models. We controlled for age and sex in Model 1 and further controlled for more covariates including ethnicity, occupation, educational level, TDI, and maternal smoking around birth in Model 2. Second, we used multiple logistic regression models to evaluate the associations between unhealthy lifestyle and frailty in two similar models. Finally, we performed a mediation analysis using R package “mediation” with 1000 simulations and documented the mediation proportions and corresponding 95% CIs in an age- and sex-adjusted model. The method for calculating the mediation proportions mediation was: average causal mediation effects/ (average causal mediation effects +average direct effects) [40].

To test the robustness of the findings, we conducted several sensitivity analyses. First, we repeated the primary analyses using another frailty measure — the frailty phenotype. Details on frailty phenotype are described in **supplemental methods**. Second, we re-estimated the associations of childhood adversity with frailty that was inputted as a continuous variable. Third, we used 0.25 as an alternative cutoff to define frailty and repeated the main analyses [3]. Fourth, we repeated the main analyses after excluding participants with missing data on lifestyles and covariates.

We used SAS version 9.4 (SAS Institute, Cary, NC) and R version 4.1.1 (2021-08-10) to conduct all analyses. P values less than 0.05 were considered statistically significant (two-tailed).

## Results

### Characteristics of participants

The characteristics of the participants are presented in **Table 1**. The age of participants was 56.4 (SD=7.7) years, and 6.6% (N=10078) of participants were defined as having frailty.

**Table 1.** Basic characteristics of study participants.

| Variables | Total<br>(N=152914) | Without frailty<br>(N=142836) | With frailty<br>(N=10078) | P value |
| --- | --- | --- | --- | --- |
| Age, years | 56.4±7.7 | 56.3±7.7 | 57.7±7.4 | <0.001 |
| Sex |  |  |  | <0.001 |
| Female | 86142 (56.3) | 80003 (56.0) | 6139 (60.9) |  |
| Male | 66772 (43.7) | 62833 (44.0) | 3939 (39.1) |  |
| Ethnicity |  |  |  | 0.004 |
| White | 148599 (97.2) | 138,823 (97.2) | 9776 (97.0) |  |
| Multiple | 798 (0.5) | 745 (0.5) | 53 (0.5) |  |
| South Asian | 1262 (0.8) | 1169 (0.8) | 93 (0.9) |  |
| Black | 1071 (0.7) | 1003 (0.7) | 68 (0.7) |  |
| Chinese | 348 (0.2) | 337 (0.2) | 11 (0.1) |  |
| Other<sup>a</sup> | 836 (0.5) | 759 (0.5) | 77 (0.8) |  |
| Educational level<sup>b</sup> |  |  |  | <0.001 |
| High | 70866 (46.3) | 67488 (47.2) | 3378 (33.5) |  |
| Intermediate | 50281 (32.9) | 46688 (32.7) | 3593 (35.7) |  |
| Low | 31767 (20.8) | 28660 (20.1) | 3107 (30.8) |  |
| Occupation |  |  |  | <0.001 |
| Working | 97657 (63.9) | 92871 (65.0) | 4786 (47.5) |  |
| Retired | 45121 (29.5) | 41542 (29.1) | 3579 (35.5) |  |
| Other | 10136 (6.6) | 8423 (5.9) | 1713 (17.0) |  |
| TDI | -1.7±2.8 | -1.8±2.8 | -1.0±3.2 | <0.001 |
| BMI, kg/m<sup>2</sup> | 26.8±4.6 | 26.6±4.4 | 29.7±6.0 | <0.001 |
| <18.5 | 857 (0.6) | 806 (0.6) | 51 (0.5) |  |
| 18.5-24.9 | 56952 (37.2) | 54882 (38.4) | 2070 (20.5) |  |
| ≥24.9 | 95105 (62.2) | 87148 (61.0) | 7957 (79.0) |  |
| Smoking, yes | 64140 (41.9) | 58885 (41.2) | 5255 (52.1) | <0.001 |
| Drinking, yes | 60764 (39.7) | 56145 (39.3) | 4619 (45.8) | <0.001 |
| Physical inactivity, yes | 39463 (25.8) | 35669 (25.0) | 3794 (37.6) | <0.001 |
| Unhealthy diet, yes | 94074 (61.5) | 87675 (61.4) | 6399 (63.5) | <0.001 |
| Maternal smoking around birth, yes | 44282 (29.0) | 40481 (28.3) | 3801 (37.7) | <0.001 |

Notes: BMI, body mass index; TDI, Townsend deprivation index.
<sup>a</sup>Other includes any races or ethnicities not otherwise specified.
<sup>b</sup>High educational level: college or university degree; Intermediate educational level: A/AS levels or equivalent, O levels/GCSEs or equivalent; Low educational level: none of the aforementioned.

### Mediation analysis of unhealthy lifestyle on associations between childhood adversity and frailty

First, **Table 2** presents the associations of childhood adversity with unhealthy lifestyle. Both individual items of childhood adversity (except for physical neglect) and cumulative childhood adversity were associated with higher unhealthy lifestyle score in the fully adjusted model (Model 2).

**Table 2.**
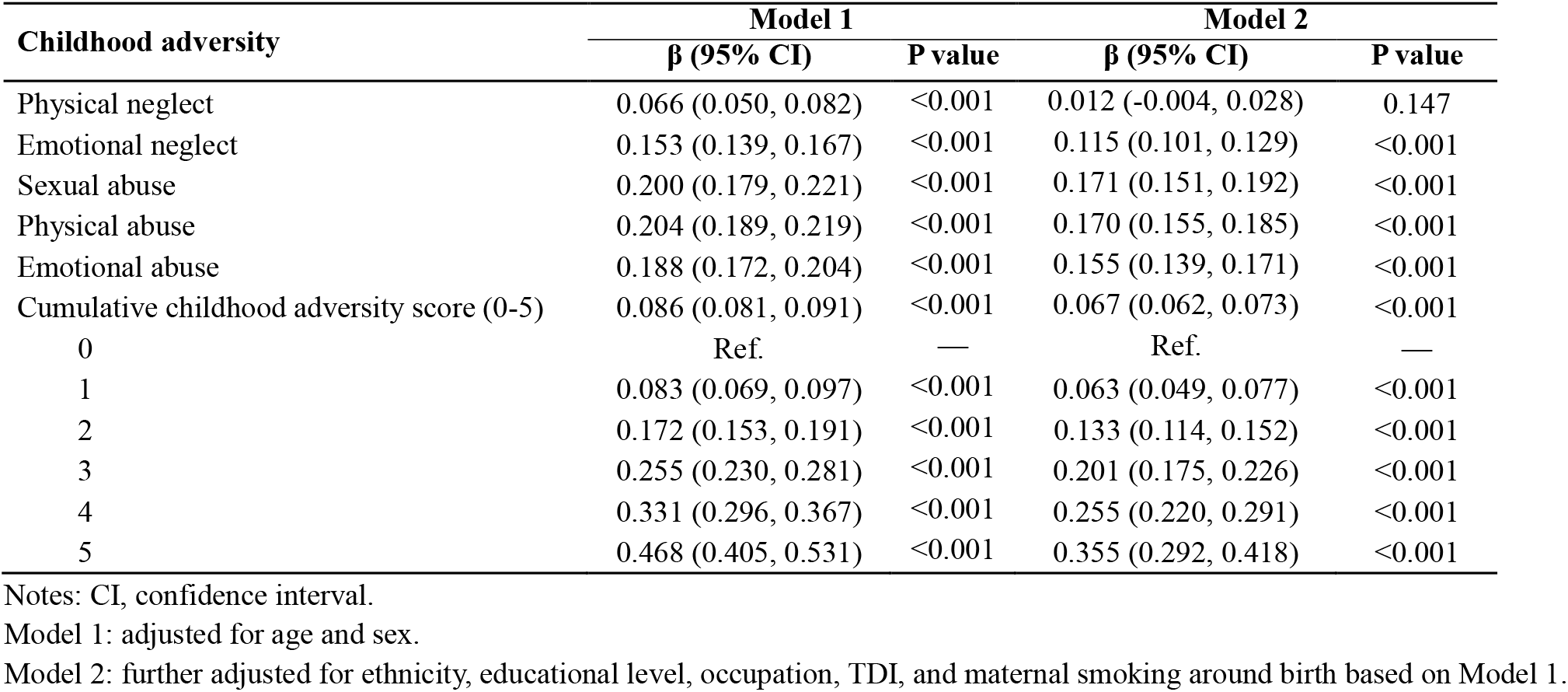
Associations of childhood adversity with unhealthy lifestyle score.

Second, **Figure S2** shows the associations of unhealthy lifestyle with frailty. In Model 1, with a one-point increase in unhealthy lifestyle score, the odds increased by 46% (OR: 1.46, 95% CI: 1.43, 1.48). In Model 2, the results remain unchanged after adjustment for other covariates.

Finally, **Figure 2** and **Figure S3** present the prevalence of frailty and frailty score by individual items and the cumulative score of childhood adversity. Compared with participants who did not experience any adversity, those who experienced adversity had a higher prevalence of frailty (P<0.001). **Table 3** shows significant associations of childhood adversity with frailty and the mediation proportion of childhood adversity in frailty attributed to unhealthy lifestyle. We found that childhood adversity was positively associated with frailty. For example, compared with participants who did not experience any adversity, those who experienced sexual abuse had increased odds (OR: 2.03; 95%CI: 1.91, 2.15) of frailty after adjustment for age and sex (Model 1). After further adjustment for ethnicity, educational level, occupation, TDI, and maternal smoking around birth, the strength of these associations decreased slightly but remained statistically significant (Model 2). After further adjusting for unhealthy lifestyle score, the associations remained robust (Model 3). For cumulative childhood adversity, a dose-response association was observed. With a one-point increase in cumulative score of childhood adversity, the odds of frailty increased by 41% (OR: 1.41; 95% CI: 1.39, 1.44) (Model 3). The mediation proportion of unhealthy lifestyle in associations between childhood adversity and frailty ranged from 4.4%-7.0%. Regarding individual items of childhood adversity, physical (mediation proportion: 8.2%) and sexual abuse (mediation proportion: 8.1%) were more likely to influence frailty through unhealthy lifestyle than other items.

**Table 3.** Associations of childhood adversity with frailty and mediation proportion of childhood adversity in frailty attributed to unhealthy lifestyle.

| Childhood adversity | Model 1 | Model 2 | Model 3 | Mediation proportion<br>(%) (95% CI)<sup>a</sup> | P value |
| --- | --- | --- | --- | --- | --- |
|  | OR (95% CI) | OR (95% CI) | OR (95% CI) |  |  |
| Physical neglect | 1.90 (1.81, 1.99) | 1.64 (1.56, 1.72) | 1.65 (1.57, 1.73) | 3.1 (2.4, 4.0) | <0.001 |
| Emotional neglect | 2.34 (2.25, 2.44) | 2.12 (2.03, 2.22) | 2.07 (1.98, 2.16) | 5.4 (4.8, 6.0) | <0.001 |
| Sexual abuse | 2.03 (1.91, 2.15) | 1.84 (1.73, 1.95) | 1.79 (1.69, 1.90) | 8.1 (7.0, 9.0) | <0.001 |
| Physical abuse | 2.14 (2.05, 2.24) | 1.96 (1.87, 2.05) | 1.89 (1.80, 1.98) | 8.2 (7.4, 9.0) | <0.001 |
| Emotional abuse | 2.76 (2.63, 2.88) | 2.48 (2.37, 2.61) | 2.41 (2.30, 2.53) | 5.1 (4.6, 6.0) | <0.001 |
| Cumulative childhood adversity score<br>(0-5) | 1.51 (1.49, 1.53) | 1.44 (1.41, 1.46) | 1.41 (1.39, 1.44) | — | — |
| 0 | Ref. | Ref. | Ref. | — | — |
| 1 | 1.60 (1.52, 1.69) | 1.53 (1.45, 1.62) | 1.52 (1.44, 1.60) | 7.0 (5.7, 9.0) | <0.001 |
| 2 | 2.39 (2.25, 2.54) | 2.18 (2.05, 2.33) | 2.13 (2.00, 2.27) | 6.6 (5.8, 7.0) | <0.001 |
| 3 | 3.48 (3.25, 3.74) | 3.03 (2.82, 3.26) | 2.92 (2.72, 3.15) | 5.8 (5.1, 7.0) | <0.001 |
| 4 | 4.96 (4.55, 5.39) | 4.02 (3.68, 4.40) | 3.88 (3.55, 4.24) | 5.0 (4.4, 6.0) | <0.001 |
| 5 | 8.04 (7.07, 9.14) | 5.85 (5.10, 6.71) | 5.49 (4.80, 6.29) | 4.4 (3.8, 5.0) | <0.001 |

Notes: OR, odds ratio; CI, confidence interval.
Model 1: adjusted for age and sex.
Model 2: further adjusted for ethnicity, educational level, occupation, TDI, and maternal smoking around birth based on Model 1.
Model 3: further adjusted for unhealthy lifestyle score based on Model 1.

**Figure 2.**
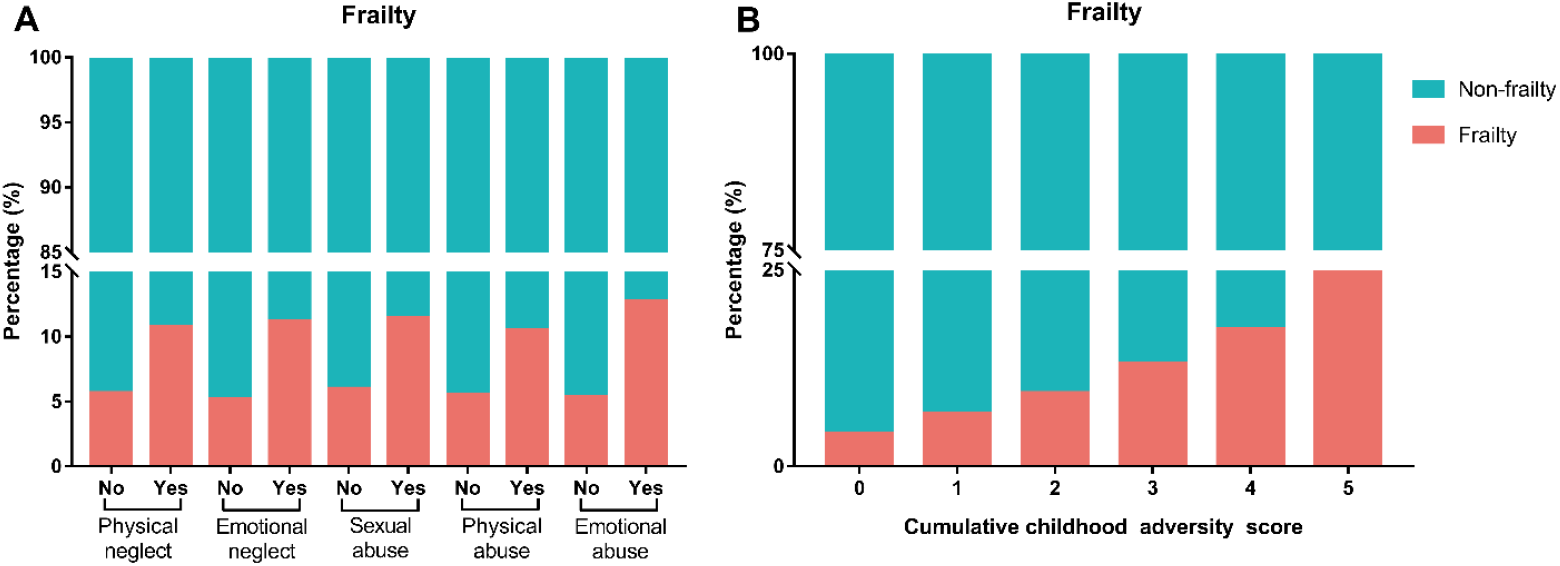
Prevalence of frailty by individual items and the cumulative score of childhood adversity. A prevalence of frailty by individual items of childhood adversity (all P<0.001); B prevalence of frailty by the cumulative score of childhood adversity (P<0.001).

### Sensitivity analyses

First, with another frailty measure (i.e., frailty phenotype), similar associations were observed with higher mediation proportion (ranging from 8.7%-17.2%) than that of frailty index (**Table S2-S4**). Second, **Table S5** shows the positive associations between cumulative childhood adversity and frailty score (β=0.010, P<0.001) in the fully adjusted model. Third, when using an alternative cutoff of 0.25 for frailty, the significant associations of childhood adversity with frailty and the mediating role of unhealthy lifestyle on the associations were maintained (**Table S6**). Fourth, after excluding participants with missing data on lifestyles, these associations remained robust (**Table S7**).

## Discussion

Our analyses of a large sample of 152914 participants of UKB aged 40-69 years, resulted in two main findings. First, we confirmed significant associations of childhood adversity with frailty, highlighting the importance of reducing traumatic experiences in early life. Second, for the first time we found that unhealthy lifestyle partially mediated the association between childhood adversity (in particular physical and sexual abuse) and frailty. The finding reveals a novel pathway and the potential of lifestyle interventions to prevent or reduce the severity of frailty among adults who have already experienced some childhood adversities.

The findings of the positive associations between childhood adversity and frailty are consistent with most previous studies [18-22]. However, one study among 182 older patients with affective disorder did not find any association of childhood adversity with frailty [23]. Our findings strengthen the associations in a large sample size of general population and have important public implications for interventions on childhood adversity in early life to diminish health inequality in late life. Frailty represents a decline of the function of multiple systems [22]. Our findings may suggest a potential of programs targeting childhood adversity in the prevention of frailty and physiological dysfunction.

To the best of our knowledge, this was the first study to demonstrate that unhealthy lifestyle partially mediated the associations of childhood adversity with frailty. Childhood adversity is linked to inadequate ability to regulate emotions and cope with stress from early-life to adulthood. As a way of managing feelings of anger and powerlessness, people who experienced childhood adversity are more likely to adopt unhealthy behaviors (e.g., smoking, drinking, unhealthy diet) [41, 42]. Frailty is widely recognized to be linked with poor diet, sedentary lifestyles, physical inactivity, and obesity [3]. Also, several studies have demonstrated that adherence to healthy lifestyle is linked to a lower risk of frailty [43-45]. These observations support our findings that childhood adversity may lead to frailty through unhealthy lifestyle. The mediating role of unhealthy lifestyle highlights the importance of modifiable lifestyle interventions to mitigate accelerated functional aging process (proportions mediation: 4.4%-8.2%) among adults who have already experienced childhood adversity. Meanwhile, the partial mediation suggests that modifiable lifestyle interventions are insufficient, and more strategies should be explored.

Given the relatively large sample, we were able to further distinguish the mediation proportions of unhealthy lifestyle in the associations between individual items of childhood adversity and frailty. Relative to other items, physical and sexual abuse were pronouncedly associated with frailty through unhealthy lifestyle. Similar to our observation of the greater associations of physical and sexual abuse with unhealthy lifestyle, previous study found that physical and sexual abuse were associated with a higher risk of smoking [46]. This provides clues that pathways from childhood adversity to health outcomes may differ by individual item of childhood adversity. A longitudinal study has indicated that the association of physical abuse with glucose metabolism may be largely explained by adulthood obesity while the association of less severe adversities (e.g., emotional neglect) was mostly explained by childhood socioeconomic factors [47]. Moreover, given the relatively small mediation proportion in our study, other mediators of biological and psychological mechanisms including adult socioeconomic status [48], attachment and mood symptoms [49] may exist. Thus, comprehensively understanding the character of diverse childhood adversities and the underlying mechanism of how they affect late-life health is necessary, and then we could tailor the programs targeting childhood adversity.

The present study has some strengths including the large sample size of middle-aged and older adults. Furthermore, a series of sensitivity analyses were performed to confirm the validity of the findings. This study also has limitations. First, the mediator (unhealthy lifestyle score) was assessed at the same time as the outcome (frailty) in the study. Longitudinal studies are needed to examine the causal relationship. Second, frailty was measured at baseline while the mental health survey was administered 6 to 10 years later, which may introduce some healthy selection. Third, recall bias was inevitable given the retrospective information of childhood adversity. However, retrospective childhood adversity records are reported to have moderate associations between childhood adversity and adverse outcomes observed in perspective-records studies [50]. Fourth, due to a lack of detailed data on childhood adversity, our study is unable to assess the association of the duration and the severity of childhood adversity with frailty. Further studies were demanded to reinforce the findings of the study. Fifth, most participants in UKB are White and have relatively high socioeconomic status [51], which may lead to selection bias and potential confounders.

## Conclusions

In this large sample of more than 150000 UK adults, childhood adversity was positively associated with frailty, and unhealthy lifestyle partially mediated the association, especially for the associations of physical and sexual abuse with frailty. Our findings highlight the potential of programs targeting on childhood adversity and lifestyle intervention strategies to promote healthy aging. A comprehensive understanding of the character of diverse childhood adversities and the underlying mechanism of how they affect late-life health is needed.

## Availability of data and materials

The datasets analysed during the current study are available in the UK Biobank repository, http://www.ukbiobank.ac.uk.

## Data Availability

All data produced in the present study are available upon reasonable request to the authors

https://www.ukbiobank.ac.uk/

## Abbreviations

UKB: UK Biobank
BMI: Body mass index
TDI: Townsend deprivation index
SD: Standard deviation
OR: Odds ratio
CI: Confidence interval

## Acknowledgments

This research has been conducted using the UK Biobank resource under application number 61856. We wish to acknowledge the UK Biobank participants who provided the sample that made the data available.

## Authors’ contributions

ZL contributed to the conception and design of the work; ZL acquired the data; GY and XC performed the analysis. All authors contributed to the interpretation of data. GY wrote the first draft of the manuscript. All authors substantively revised the manuscript, read and approved the final manuscript.

## Ethics approval and consent to participate

The UK Biobank study was approved by the North West Multi-Centre Research Ethics Committee (11/NW/0382) and written informed consent was provided by each participant before the study. UK Biobank also has approval from the North West Multi-Centre Research Ethics Committee as a Research Tissue Bank (RTB) approval in 2011 and is renewed every 5 years. According to RTB approval, researchers are allowed to use data from UK Biobank without an additional ethical clearance.

## Consent for publication

Not applicable.

