## Supplementary material for "Association of childhood adversity with frailty and the mediating role of unhealthy lifestyle: Findings from the UK biobank"

**Supplement Methods**

**Another frailty measure (frailty phenotype)**

Frailty was measured by five phenotype items as follows: 1) for weight loss: “Compared with one year ago, has your weight changed?” with coded 1 for “lose weight” and 0 for other answers; 2) for exhaustion: “Over the past two weeks, how often have you felt tired or had little energy?” with code 1 for “more than half the day” or “nearly every day”, and 0 for others; 3) for slow walking speed: “How would you describe your usual walking pace?” with coded 1 for “slow” and 0 for others; 4) for low physical activity: “In the last 4 weeks did you spend any time doing the following? (You can select more than one answer)” with coded 1 for “None or light activity with a frequency of once per week or less” and 0 for “medium or heavy activity, or light activity more than once per week”; 5) for low grip strength: measured grip strength (cutoffs adjusted based on sex and body mass index) [1]. The summary score of 5 items valued 0-2 was categorized as non-frailty, while valued ≥3 was categorized as frailty [2].

**Table S1. Assessment of childhood adversity.**

| **Childhood adversityquestions^a^** | | **Responses/description** | **Cut off [3]** |
| --- | --- | --- | --- |
| Physical neglect | Someone to take to doctor when needed as a child. | 0, Never true; 1, Rarely true; 2, Sometimes true; 3, Often; 4, Very often true | ≤3 |
| Emotional neglect | Felt loved as a child. | 0, Never true; 1, Rarely true; 2, Sometimes true; 3, Often; 4, Very often true | ≤2 |
| Sexual abuse | Sexually molested as a child. | 0, Never true; 1, Rarely true; 2, Sometimes true; 3, Often; 4, Very often true | ≥1 |
| Physical abuse | Physically abused by family as a child. | 0, Never true; 1, Rarely true; 2, Sometimes true; 3, Often; 4, Very often true | ≥1 |
| Emotional abuse | Felt hated by family member as a child. | 0, Never true; 1, Rarely true; 2, Sometimes true; 3, Often; 4, Very often true | ≥1 |
| Cumulative childhood adversity risk | Summary score of childhood adversity | Summary score of five items (0-5). | — |

Notes: ^a^All items were referred to as, “When I was growing up (age <16 years old)”.

**Table S2.** **Basic characteristics of study participants by frailty phenotype.**

| **Variables** | **Total**  **(N=152914)** | **Without frailty (N=149388)** | **With frailty (N=3526)** | **P value** |
| --- | --- | --- | --- | --- |
| Age, years | 56.4±7.7 | 56.3±7.7 | 56.9±7.4 | <0.001 |
| Sex |  |  |  | <0.001 |
| Female | 86142 (56.3) | 83770 (56.1) | 2372 (67.3) |  |
| Male | 66772 (43.7) | 65618 (43.9) | 1154 (32.7) |  |
| Ethnicity |  |  |  | <0.001 |
| White | 148599 (97.2) | 145254 (97.2) | 3345 (94.9) |  |
| Multiple | 798 (0.5) | 766 (0.5) | 32 (0.9) |  |
| South Asian | 1262 (0.8) | 1201 (0.8) | 61 (1.7) |  |
| Black | 1071 (0.7) | 1029 (0.7) | 42 (1.2) |  |
| Chinese | 348 (0.2) | 343 (0.2) | 5 (0.1) |  |
| Other^a^ | 836 (0.5) | 795 (0.5) | 41 (1.2) |  |
| Educational level^b^ |  |  |  | <0.001 |
| High | 70866 (46.3) | 69701 (46.7) | 1165 (33.0) |  |
| Intermediate | 50281 (32.9) | 48993 (32.8) | 1288 (36.5) |  |
| Low | 31767 (20.8) | 30694 (20.5) | 1073 (30.4) |  |
| Occupation |  |  |  | <0.001 |
| Working | 97657 (63.9) | 95952 (64.2) | 1705 (48.4) |  |
| Retired | 45121 (29.5) | 44068 (29.5) | 1053 (29.9) |  |
| Other | 10136 (6.6) | 9368 (6.3) | 768 (21.8) |  |
| TDI | -1.7±2.8 | -1.7±2.8 | -0.7±3.3 | <0.001 |
| BMI, kg/m^2^ | 26.8±4.6 | 26.7±4.5 | 30.9±6.7 | <0.001 |
| <18.5 | 857 (0.6) | 833 (0.6) | 24 (0.7) |  |
| 18.5-24.9 | 56952 (37.2) | 56359 (37.7) | 593 (16.8) |  |
| ≥24.9 | 95105 (62.2) | 92196 (61.7) | 2909 (82.5) |  |
| Smoking, yes | 64140 (41.9) | 62444 (41.8) | 1696 (48.1) | <0.001 |
| Drinking, yes | 60764 (39.7) | 59168 (39.6) | 1596 (45.3) | <0.001 |
| Physical inactivity, yes | 39463 (25.8) | 37359 (25.0) | 2104 (59.7) | <0.001 |
| Unhealthy diet, yes | 94074 (61.5) | 91730 (61.4) | 2344 (66.5) | <0.001 |
| Maternal smoking around birth, yes | 44282 (29.0) | 43008 (28.8) | 1274 (36.1) | <0.001 |

Notes: BMI, body mass index; TDI, Townsend deprivation index.

^a^Other includes any races or ethnicities not otherwise specified.

^b^High educational level: college or university degree; Intermediate educational level: A/AS levels or equivalent, O levels/GCSEs or equivalent; Low educational level: none of the aforementioned.

**Table S3. Associations of childhood adversity with frailty phenotype and mediation proportion of childhood adversity in frailty phenotype attributed to unhealthy lifestyle.**

| **Childhood adversity** | **Model 1** | **Model 2** | **Model 3** | **Mediation proportion**  **(%) (95% CI)^a^** | **P value** |
| --- | --- | --- | --- | --- | --- |
|  | **OR (95% CI)** | **OR (95% CI)** | **OR (95% CI)** |  |  |
| Physical neglect | 1.70 (1.57, 1.84) | 1.40 (1.30, 1.52) | 1.40 (1.29, 1.52) | 5.7 (4.2, 7.0) | <0.001 |
| Emotional neglect | 1.82 (1.70, 1.95) | 1.59 (1.47, 1.70) | 1.51 (1.40, 1.62) | 11.9 (10.2, 14.0) | <0.001 |
| Sexual abuse | 1.72 (1.56, 1.89) | 1.51 (1.37, 1.67) | 1.40 (1.27, 1.55) | 15.7 (12.6, 19.0) | <0.001 |
| Physical abuse | 1.80 (1.67, 1.94) | 1.59 (1.47, 1.72) | 1.47 (1.36, 1.59) | 16.4 (14.1, 19.0) | <0.001 |
| Emotional abuse | 2.03 (1.88, 2.19) | 1.73 (1.60, 1.88) | 1.62 (1.49, 1.75) | 11.4 (9.8, 13.0) | <0.001 |
| Cumulative childhood adversity score (0-5) | 1.35 (1.32, 1.38) | 1.24 (1.21, 1.27) | 1.21 (1.18, 1.24) | — | — |
| 0 | Ref. | Ref. | Ref. | — | — |
| 1 | 1.34 (1.23, 1.46) | 1.25 (1.15, 1.36) | 1.23 (1.12, 1.34) | 17.2 (12.5, 26.0) | <0.001 |
| 2 | 1.86 (1.68, 2.05) | 1.64 (1.48, 1.82) | 1.55 (1.40, 1.72) | 14.3 (11.8, 18.0) | <0.001 |
| 3 | 2.28 (2.02, 2.58) | 1.88 (1.66, 2.13) | 1.73 (1.53, 1.96) | 14.2 (11.7, 17.0) | <0.001 |
| 4 | 3.38 (2.94, 3.88) | 2.54 (2.20, 2.93) | 2.28 (1.97, 2.63) | 10.0 (8.5, 12.0) | <0.001 |
| 5 | 4.80 (3.92, 5.89) | 3.01 (2.44, 3.72) | 2.62 (2.12, 3.25) | 8.7 (7.3, 10.0) | <0.001 |

Notes: OR, odds ratio; CI, confidence interval.
Model 1: adjusted for age and sex.
Model 2: further adjusted for ethnicity, educational level, occupation, Townsend deprivation index, and maternal smoking around birth based on Model 1.

Model 3: further adjusted for unhealthy lifestyle score based on Model 2.

^a^The model included age, sex, and unhealthy lifestyle score.

**Table S4. Associations of unhealthy lifestyle with frailty phenotype.**

|  | **Model 1** | | **Model 2** | |
| --- | --- | --- | --- | --- |
|  | **OR (95% CI)** | **P value** | **OR (95% CI)** | **P value** |
| Unhealthy lifestyle score | 1.74 (1.69, 1.80) | <0.001 | 1.65 (1.61, 1.71) | <0.001 |

Notes: OR, odds ratio; CI, confidence interval.

Model 1: adjusted for age and sex.

Model 2: further adjusted for ethnicity, educational level, occupation, Townsend deprivation index, and maternal smoking around birth based on Model 1.

**Table S5. Associations between childhood adversity** **and frailty score.**

| **Childhood adversity** | **Model 1** | | **Model 2** | | **Model 3** | |
| --- | --- | --- | --- | --- | --- | --- |
|  | **β (SE)** | **P value** | **β (SE)** | **P value** | **β (SE)** | **P value** |
| Physical neglect | 0.016 (0.0005) | <0.001 | 0.012 (0.0005) | <0.001 | 0.012 (0.0005) | <0.001 |
| Emotional neglect | 0.024 (0.0004) | <0.001 | 0.022 (0.0004) | <0.001 | 0.021 (0.0004) | <0.001 |
| Sexual abuse | 0.021 (0.0006) | <0.001 | 0.019 (0.0006) | <0.001 | 0.018 (0.0006) | <0.001 |
| Physical abuse | 0.020 (0.0005) | <0.001 | 0.018 (0.0005) | <0.001 | 0.017 (0.0004) | <0.001 |
| Emotional abuse | 0.029 (0.0005) | <0.001 | 0.027 (0.0005) | <0.001 | 0.025 (0.0005) | <0.001 |
| Cumulative childhood adversity score (0-5) | 0.012 (0.0002) | <0.001 | 0.011 (0.0002) | <0.001 | 0.010 (0.0002) | <0.001 |

Notes: SE, standard error.

Model 1: adjusted for age and sex.
Model 2: further adjusted for ethnicity, educational level, occupation, Townsend deprivation index, and maternal smoking around birth based on Model 1.

Model 3: further adjusted for unhealthy lifestyle score based on Model 2.

**Table S6.** **Associations of childhood adversity with frailty (cut-off: 0.25) and mediation proportion of childhood adversity in frailty (cut-off: 0.25) attributed to unhealthy lifestyle.**

| **Childhood adversity** | **Model 1** | **Mediation proportion**  **(%) (95% CI)^a^** | **P value** |
| --- | --- | --- | --- |
|  | **OR (95% CI)** |  |  |
| Physical neglect | 1.78 (1.65, 1.92) | 3.2 (2.4, 4.0) | <0.001 |
| Emotional neglect | 2.20 (2.05, 2.36) | 5.7 (5.0, 7.0) | <0.001 |
| Sexual abuse | 2.01 (1.83, 2.20) | 7.1 (6.1, 8.0) | <0.001 |
| Physical abuse | 2.06 (1.91, 2.21) | 7.8 (6.9, 9.0) | <0.001 |
| Emotional abuse | 2.73 (2.54, 2.94) | 4.9 (4.4, 6.0) | <0.001 |
| Cumulative childhood adversity score (0-5) | 1.46 (1.43, 1.50) | — | — |
| 0 | Ref. | — | — |
| 1 | 1.60 (1.46, 1.75) | 7.4 (5.5, 10.0) | <0.001 |
| 2 | 2.27 (2.05, 2.52) | 7.3 (6.1, 9.0) | <0.001 |
| 3 | 3.52 (3.14, 3.94) | 5.6 (4.9, 6.0) | <0.001 |
| 4 | 4.31 (3.76, 4.94) | 5.0 (4.3, 6.0) | <0.001 |
| 5 | 6.61 (5.47, 7.99) | 3.7 (3.1, 4.0) | <0.001 |

Notes: OR, odds ratio; CI, confidence interval.

Model 1: adjusted for age, sex, ethnicity, educational level, occupation, Townsend deprivation index, maternal smoking around birth and unhealthy lifestyle score.

^a^The model included age, sex, and unhealthy lifestyle score.

**Table S7. Associations of childhood adversity with frailty and mediation proportion of childhood adversity in frailty attributed to unhealthy lifestyle after excluding participants with missing data on lifestyles and covariates (N=113950).**

| **Childhood adversity** | **Model 1** | **Model 2** | **Model 3** | **Mediation proportion**  **(%) (95% CI)^a^** | **P value** |
| --- | --- | --- | --- | --- | --- |
|  | **OR (95% CI)** | **OR (95% CI)** | **OR (95% CI)** |  |  |
| Physical neglect | 1.86 (1.75, 1.97) | 1.63 (1.53, 1.73) | 1.63 (1.54, 1.73) | 2.6 (1.8, 4.0) | <0.001 |
| Emotional neglect | 2.34 (2.22, 2.47) | 2.13 (2.02, 2.25) | 2.07 (1.97, 2.19) | 5.1 (4.4, 6.0) | <0.001 |
| Sexual abuse | 1.97 (1.83, 2.11) | 1.85 (1.72, 1.99) | 1.76 (1.64, 1.90) | 8.1 (6.8, 9.0) | <0.001 |
| Physical abuse | 2.14 (2.02, 2.26) | 1.98 (1.87, 2.09) | 1.89 (1.78, 2.00) | 7.9 (7.0, 9.0) | <0.001 |
| Emotional abuse | 2.75 (2.61, 2.92) | 2.52 (2.38, 2.67) | 2.43 (2.29, 2.57) | 4.9 (4.3, 6.0) | <0.001 |
| Cumulative childhood adversity score (0-5) | 1.51 (1.48, 1.54) | 1.44 (1.41, 1.47) | 1.42 (1.39, 1.44) | — | — |
| 0 | Ref. | Ref. | Ref. | — | — |
| 1 | 1.63 (1.53, 1.74) | 1.57 (1.47, 1.67) | 1.55 (1.45, 1.65) | 6.4 (5.2, 8.0) | <0.001 |
| 2 | 2.36 (2.19, 2.55) | 2.17 (2.02, 2.35) | 2.10 (1.95, 2.27) | 6.7 (6.0, 8.0) | <0.001 |
| 3 | 3.58 (3.29, 3.91) | 3.14 (2.88, 3.43) | 3.01 (2.75, 3.29) | 5.1 (4.3, 6.0) | <0.001 |
| 4 | 4.94 (4.44, 5.49) | 4.17 (3.74, 4.65) | 3.93 (3.52, 4.39) | 4.8 (4.1, 6.0) | <0.001 |
| 5 | 8.32 (7.05, 9.81) | 6.19 (5.21, 7.36) | 5.65 (4.75, 6.73) | 4.4 (3.6, 5.0) | <0.001 |

Notes: OR, odds ratio; CI, confidence interval.
Model 1: adjusted for age and sex.
Model 2: further adjusted for ethnicity, educational level, occupation, Townsend deprivation index, and maternal smoking around birth based on Model 1.

Model 3: further adjusted for unhealthy lifestyle score based on Model 2.

^a^The model included age, sex, and unhealthy lifestyle score.

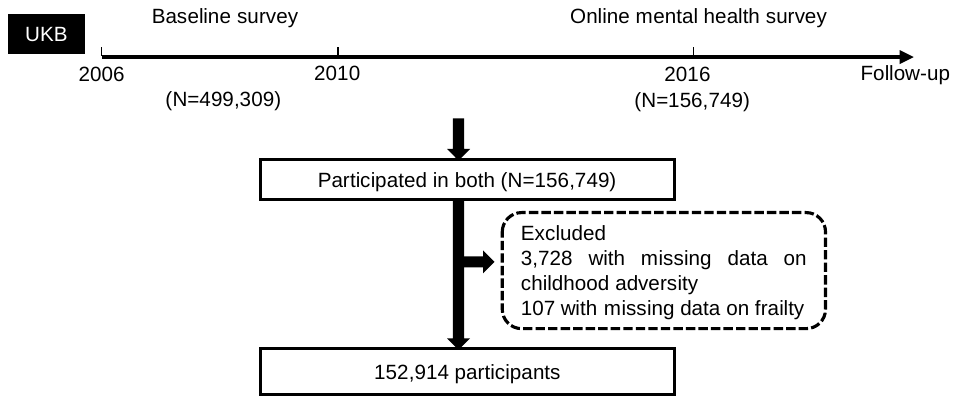

**Figure S1. Flow chart of the analytic sample.**

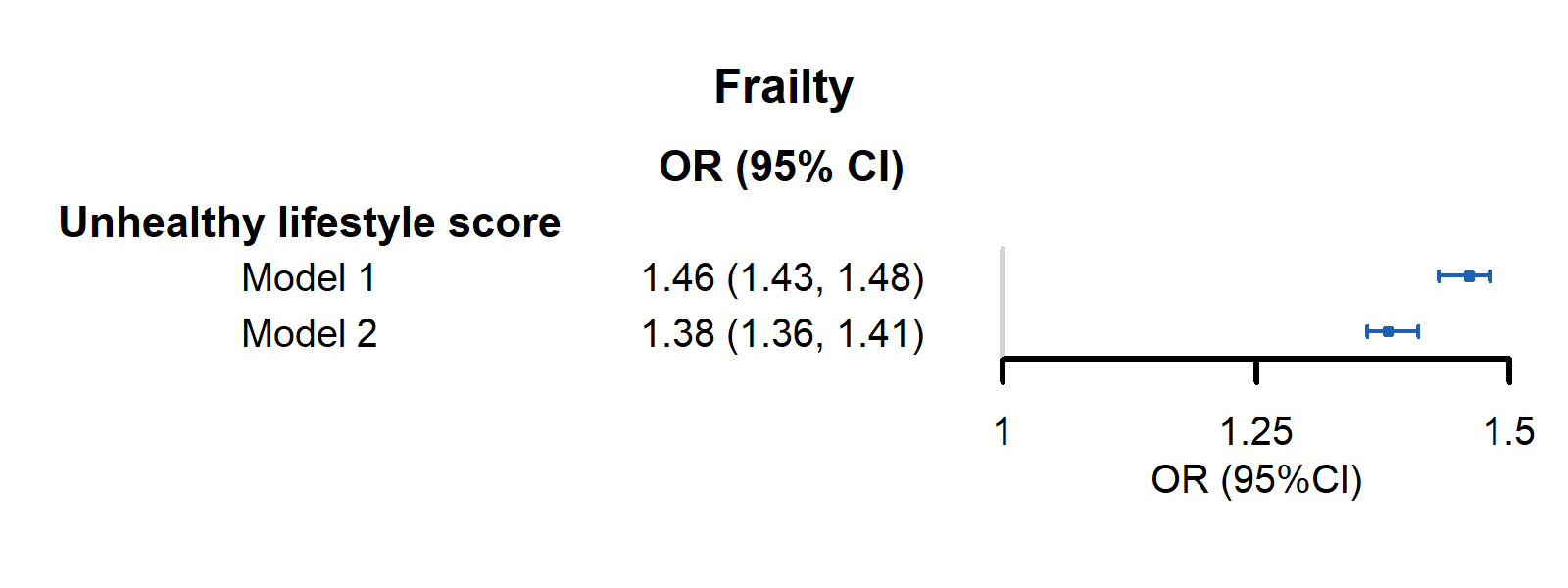
**Figure S2. The associations between unhealthy lifestyle score and frailty (one-point increase in unhealthy lifestyle score).**

Notes: OR, odds ratio; CI, confidence interval.

Model 1: adjusted for age and sex.

Model 2: further adjusted for ethnicity, educational level, occupation, Townsend deprivation index, and maternal smoking around birth based on Model 1.

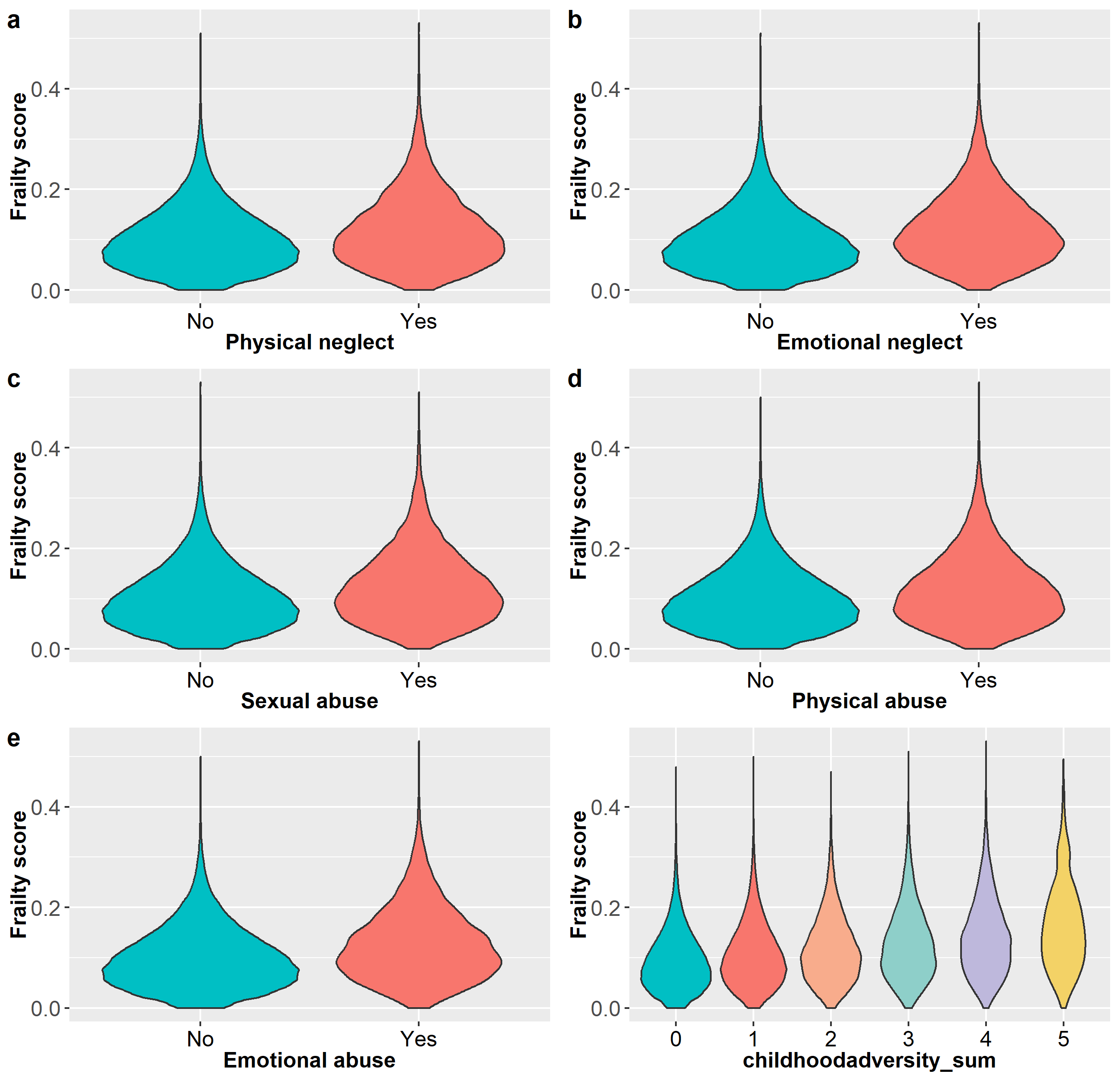

**Figure S3. Violin plot of frailty score by individual items and the cumulative score of childhood adversity.** Frailty score by **A** physical neglect, **B** emotional neglect, **C** sexual abuse, **D** physical abuse, **E** emotional abuse, and **F** the cumulative score of childhood adversity.
